# Frontal tDCS reduces alcohol relapse rates by increasing connections from left dorsolateral prefrontal cortex to addiction networks

**DOI:** 10.1101/2022.11.28.22282521

**Authors:** Jazmin Camchong, Mark Fiecas, Casey S. Gilmore, Matt Kushner, Erich Kummerfeld, Bryon A. Mueller, Donovan Roediger, Kelvin O. Lim

**Affiliations:** University of Minnesota Department of Psychiatry and Behavioral Sciences, 2312 S. 6th St., Floor 2, Suite F-275, Minneapolis, MN 55454, United States of America; University of Minnesota School of Public Health, 420 Delaware St SE, Minneapolis, MN 55455, United States of America; Minneapolis VA Health Care System, 1 Veterans Dr., Minneapolis, MN 55417, United States of America; University of Minnesota Institute for Health Informatics, 8-100 Phillips-Wangensteen Building, 516 Delaware Street SE, Minneapolis, MN 55455, United States of America

**Keywords:** Alcohol use disorder, transcranial direct current stimulation, causal connectivity, dorsolateral prefrontal cortex, incentive salience, relapse

## Abstract

**Background:** Brain-based interventions are needed to address persistent relapse in alcohol use disorder (AUD). Neuroimaging evidence suggests higher fronto-striatal connectivity as well as higher within-network connectivity of theoretically defined addiction networks is associated with reduced relapse rates and extended abstinence during follow-up periods.

**Objective/Hypothesis:** A longitudinal randomized double-blind sham-controlled clinical trial investigated whether a non-invasive neuromodulation intervention delivered during early abstinence can (i) modulate connectivity of addiction networks supporting abstinence and (ii) improve relapse rates. Hypotheses: Active transcranial direct current stimulation (tDCS) will (i) increase connectivity of addiction networks known to support abstinence and (ii) reduce relapse rates.

**Methods:** Short-term abstinent AUD participants (n=60) were assigned to 5 days of either active tDCS or sham during cognitive training. Causal discovery analysis (CDA) examined the directional influence from left dorsolateral prefrontal cortex (LDLPFC, stimulation site) to addiction networks that support abstinence.

**Results:** Active tDCS had an effect on the average strength of CDA-determined connectivity from LDLPFC to the incentive salience and negative emotionality addiction networks - increasing in the active tDCS group and decreasing in the sham group. Active tDCS had an effect on relapse rates 1-month following the intervention, with lower probability of relapse in the active tDCS vs. sham groups. Active tDCS showed an unexpected sex-dependent effect on relapse rates.

**Conclusion:** Our results suggest that LDLPFC stimulation delivered during early abstinence has an effect on addiction networks supporting abstinence and on relapse rates. The unexpected sex-dependent neuromodulation effects need to be further examined in larger clinical trials.

## INTRODUCTION

Alcohol use disorder (AUD) continues to directly afflict about 24 million individuals and impact the lives of many millions more. The low success rate of current psychosocially-based (e.g. 12-step) treatment programs (∼64% relapse within a year) highlights the need for novel and effective interventions that target underlying biological and cognitive factors contributing to relapse. Non-invasive neuromodulation interventions are continuing to provide promising results in AUD [1–3].

Neuroimaging research has provided key evidence by identifying targets for *brain-based* non-invasive interventions. Because of the involvement of frontal-striatal networks in executive functioning and reward processing, mounting evidence suggests that higher resting state functional connectivity (RSFC) of these networks during early abstinence supports abstinence maintenance. Our past resting-state fMRI studies contributed to this literature by reporting that RSFC between dorsolateral prefrontal cortex (DLPFC) and nucleus accumbens (NAcc) during early abstinence is associated with future treatment outcome. Specifically: (i) AUD participants with long-term abstinence have higher DLPFC-NAcc RSFC vs. those with short-term abstinence or controls [4,5]; (ii) Reduced DLPFC-NAcc RSFC during early abstinence can predict subsequent relapse and faster time to relapse [6,7]; and (iii) DLPFC-NAcc RSFC decreases during early abstinence among those that subsequently relapse but not in those that remained abstinent [8,9]. Our findings are consistent with the neuroimaging literature in other substance use disorders (e.g. nicotine, stimulant use disorder) that propose that higher RSFC between DLPFC and brain regions known to process reward can protect from relapse risk [10–13].

Adopting a more comprehensive approach that extends beyond regions of interest, we examined RSFC within theoretically defined addiction networks: the incentive salience (IS), negative emotionality (NE), executive functioning Go (EF/Go), and executive functioning Stop (EF/Stop) networks [14,15]. Our most recent publication [16] demonstrated that the strength of RSFC within four addiction networks measured during early abstinence was associated with abstinence maintenance, while RSFC within a sensory processing -visual-network was not [16]. The addiction network that showed the strongest effect was the IS; higher within-network RSFC in IS during early abstinence decreased the odds of relapse in the subsequent month [16].

The above data indicates that early abstinence is a critical period during which enhanced RSFC of frontal and addiction networks is a protective factor that supports subsequent abstinence. This evidence suggests that non-invasive neuromodulation interventions that support abstinence maintenance need to be designed to increase DLPFC RSFC and addiction network RSFC during early abstinence.

### Effect of DLPFC stimulation on treatment outcome in substance use disorder

Preclinical and clinical studies suggest DLPFC stimulation is a promising intervention target for neuromodulation trials in substance use disorder. Preclinical research has reported reduced motivation to consume alcohol, reduced alcohol consumption and reduced reacquisition of ethanol consumption after multiple sessions of transcranial direct current stimulation (tDCS) to the frontal cortex in murine models [17,18]. Clinical studies have reported reduced craving and risk of relapse after multiple sessions (five [2,3] or ten sessions [19]) of DLPFC stimulation.

While a significant amount of literature suggests promising effects of DLPFC stimulation with tDCS [1] and other non-invasive stimulation methods [20–24] on treatment outcomes, inconsistent findings still remain. There are reports of (i) no additive effects of tDCS on self-reported craving in individuals with AUD undergoing mindfulness-based relapse prevention therapy (when administered once a week for eight weeks[25]) and (ii) no effects of tDCS (after four consecutive sessions) on the amount of alcohol consumption (number of drinks or percent of drinking days) in risk-drinkers undergoing cognitive bias modification [26]. Inconsistent findings of tDCS’s effect on treatment outcome could be related to infrequent spacing of intervention sessions [25], studying an at risk population rather than individuals with chronic alcohol use with diagnosed AUD [26], or administering tDCS with concurrent tasks not designed to enhance executive functioning [25,26]. The present study administered five consecutive days of DLPFC stimulation sessions on short-term abstinent individuals diagnosed with AUD using a combination of tDCS and concurrent cognitive training.

### Effect of DLPFC stimulation on RSFC

There is growing evidence from non-invasive intervention studies that DLPFC stimulation can modulate RSFC [27–31]. An early study on healthy controls, reported that one tDCS session stimulating bilateral DLPFC modulated RSFC of the default mode, fronto-parietal, and interceptive networks [32]. Studies in individuals with substance use disorder continue to add consistent findings on the effect of DLPFC stimulation on RSFC. For example, a study on abstinent individuals with methamphetamine use disorder, reported that one tDCS session stimulating bilateral DLPFC modulated RSFC of the default mode, executive functioning, and incentive salience networks [33]. Another study on individuals with AUD reported that five tDCS sessions stimulating DLPFC produced a significant increase in global efficiency and a significant reduction in global clustering; network-based statistical analysis identified a significant increase in RSFC of a specific network involving prefrontal cortex in individuals with AUD [2]. Most recently, a study on individuals with gambling disorder reported that a single session of DLPFC stimulation increased RSFC between DLPFC and the right superior parietal lobule [34]. While previous studies in substance use disorder have examined RSFC between DLPFC and either regions of interest (e.g. nucleus accumbens, default mode network, parietal cortex) or executive control networks, it is clinically important to determine whether tDCS modulates RSFC between DLPFC and theoretically defined addiction networks [14,15]. The current study is the first to use a non-invasive neurostimulation intervention delivered during early abstinence in AUD to investigate the directional effects of DLPFC stimulation on RSFC of theoretically defined addiction networks known to support abstinence [16].

### From RSFC to resting state causal connectivity

Our past studies have been key in identifying RSFC-based markers of extended abstinence and relapse. RSFC, however, is limited because it lacks information on the directionality of identified significant correlations. New methods have been recently developed for fMRI to determine not only the strength and sign (positive or negative) of significant correlations, but also their directionality, allowing the identification of causal relationships. This analysis methodology was used in the present study because it allows us to determine the directional influence of DLPFC stimulation on RSFC within addiction networks known to support abstinence.

Causal discovery analysis (CDA) is a powerful data-driven methodology that allows the determination of the directed influence (i.e. the causal connectivity) between variables of interest. A large-scale AUD study [35] used CDA to determine the direct causal influence between resting state networks, phenotypic domains, and AUD symptom severity. Rawls et al (2021) reported a learned causal model identifying a resting state network whose functional connectivity had the most directional influence on AUD symptom severity, a prefrontal cortex network mediating executive functioning (e.g. inhibitory control, working memory). A following paper by the same group [36] detailed how CDA can be used to determine whole-brain causal connectomes from rest fMRI data determining (i) edges or connections between regions and (ii) edge orientation [37,38]. In the current study, we applied this CDA methodology [36] to determine whether the causal relationship between the left dorsolateral prefrontal cortex (LDLPFC, tDCS stimulation site [39]) and theoretically defined addiction networks known to support abstinence [16] changes following intervention.

Taken together, the present study adds evidence to the neuromodulation clinical trials in addiction by combining: (i) The use of tDCS paired with cognitive training to engage executive control during the stimulation session; (ii) Tracking relapse outcome over a 4-month follow-up period, longer than follow-up periods in previous clinical trials [3,40]; (iii) Collecting pre- and post-tDCS resting fMRI data, and (iv) Using CDA to determine the directionality of the standard correlational associations to be able to draw causal associations [35] between the stimulation site (LDLPFC) and theoretically defined addiction networks known to support abstinence [14,16].

## MATERIALS AND METHODS

### Participants

A total of 81 participants with AUD were consented (Inclusion/Exclusion criteria in **Supplementary Material A**). All participants were recruited during early abstinence, 1-2 weeks after being admitted to a 28-day inpatient addiction treatment program in Minneapolis, MN (number of days abstinent until MRI session M=24.33, SD=16.47). All participants provided written informed consent and received monetary compensation for participating. The consent process and all procedures were approved by the Institutional Review Board at the University of Minnesota.

From the 81 participants, 21 did not have complete fMRI (functional magnetic resonance imaging) data because: 10 left the treatment program and were no longer reachable (9 after their pre-intervention neuroimaging session, 1 after the first tDCS session); 4 were found to be no longer eligible (1 because of identified cognitive impairment, 2 because the identified *primary* substance use disorder diagnosis was not alcohol, but stimulant and heroin and 1 because of identified unknown metal in their bodies), 3 voluntarily withdrew participation before the initial tDCS session, 3 were excluded from group analyses because their fMRI data did not meet our image quality threshold (Resting State Data Quality Assessment in **Supplementary Material C**), 1 because of scanner hardware issues during scan. As a result, pre- and post-intervention fMRI data was available for 60 participants (**Table 1**). Timeline Follow-Back (TLFB) [41] was used to record alcohol/drug use history for the past 6 months before entering the treatment program (**Table 1**).

**Table 1.** Demographics and history of alcohol use in participants who had complete fMRI data.

| Characteristic | Intervention Group |  |  |  |
| --- | --- | --- | --- | --- |
|  | All AUD | Active tDCS | Sham | Active tDCS vs. Sham |
|  | (n=60) | (n=31) | (n=29) |  |
|  | Mean or *n* | Mean or *n* | Mean or *n* | T-test or |
| | (SD or %) | (SD or %) | (SD or %) | $\chi^2$ (italics) |
| Age | 41.65 (9.60) | 39.84 (9.97) | 43.59 (8.94) | p=0.13 |
| Years of Education | 14.32 (1.98) | 14.72 (2.10) | 13.89 (1.78) | p=0.12 |
| Female, n (%) | 21 (35.0%) | 11 (52.4%) | 10 (47.6%) | p=0.94 |
| Age of AUD onset | 28.10 (9.52) | 26.80 (7.23) | 29.54 (11.52) | p=0.28 |
| # of standard drinks: Past 6-months | 2642.00 (2192.62) | 2714.03 (1949.76) | 2569.97 (2451.90) | p=0.823 |
| # of drinking days: Past 6-months | 100.11 (61.40) | 101.26 (60.48) | 98.96 (63.59) | p=0.898 |
| # of days abstinent until the baseline MRI session | 24.33 (16.47) | 25.08 (15.93) | 23.58 (17.30) | p=0.756 |

AUD, Alcohol Use Disorder; MRI, Magnetic Resonance Imaging; SD, Standard Deviation; tDCS, transcranial direct current stimulation. $\chi^2$ , Chi-Square; p, significance probability value.

### Intervention

#### Transcranial Direct Current Stimulation (tDCS)

tDCS was performed with the StarStim wireless neurostimulator system (Neuroelectrics, Inc., Barcelona, Spain). Direct current was induced by two circular rubber carbon core electrodes in saline-soaked surface sponges (25 cm^2^), placed in a neoprene headcap with marked locations based on the 10-20 EEG system [39]. The anodal electrode was at F3 and the cathodal electrode was at F4. Intervention sessions took place twice per day (13 minute duration each, separated by 20 minutes) on five consecutive days [42,43]. For active stimulation, participants received a constant current of 2 mA intensity for 13 min (30 seconds ramp up/down). For sham stimulation, current was ramped down (30 s) immediately after the initial ramp up period, and then ramped up (30 s) right before the final ramp down portion of the session. Participants were administered a questionnaire before and after each tDCS session assessing the presence and severity of potential side effects^1^.

#### Cognitive task completed during tDCS

Preclinical and clinical literature suggests that chronic substance use is associated with poor cognitive flexibility as measured by the reversal learning set-shifting task [44–47]. We administered the 4-choice reversal learning task [48](**Supplementary Material B**) concurrently with each tDCS. Task administration started after the 30 seconds tDCS ramp up.

#### Relapse Metrics

All participants were abstinent at the time of the pre- and post-intervention MRI and neuromodulation sessions because these sessions were completed when they were inpatients in the addiction treatment program. Participants underwent random alcohol/drug tests in the treatment program. After participants were discharged from the addiction treatment program they completed in-person interviews at the 1- and 4-month follow-up timepoints. Participants were in the relapsing group (REL) if they reported consuming at least one drink. Participants who had not consumed any alcohol/drug were considered to be in the abstaining group (ABS). One- and 4-month relapse outcomes were defined as separate outcome variables to be able to examine potential durability intervention effects.

#### Brain Imaging Metrics

Brain imaging data acquisition, quality assessment, preprocessing and individual level analyses are in **Supplementary Material C**.

#### Group analyses to determine intervention effects

To determine intervention effects on causal connectivity, a 2 (Intervention: Active tDCS vs. sham) x 2 (Timepoint: pre-intervention vs. post-intervention fMRI) general linear model correcting for baseline was conducted with causal connectivity (standardized r score) between LDLPFC and each addiction network as the dependent variable.

To determine whether causal connectivity changes were different depending on treatment outcome, a 2 (Outcome: Relapsed vs. Abstained) x 2 (Timepoint: pre-intervention vs. post-intervention fMRI) general linear model correcting for baseline was conducted with causal connectivity (standardized r score) between LDLPFC and each addiction network as the dependent variable.

To determine the intervention effects on relapse outcome, a Pearson Chi square test was conducted with relapse outcome (relapsed vs. abstained) as the dependent variable and intervention (active tDCS vs. sham) as the independent variable. Additionally, because Cox-proportional hazards regression revealed that women had higher likelihood of relapse than men (**Supplementary Material F**), the same analysis was conducted with the sample split by sex as a biological variable.

To determine whether the change in causal connectivity as a result of the intervention is associated with relapse outcome, a logistic regression was conducted with relapse outcome as the dependent variable, intervention type as the between-groups factor, and change in causal connectivity from LDLPFC to each addiction network as a covariate.

## RESULTS

### Demographic, clinical and behavioral comparison between groups

There were no significant differences in age, education, number of women, age of AUD onset, AUD severity in the past six months, or length of abstinence before the baseline MRI session between intervention groups (Active tDCS vs. Sham)(**Table 1**). There were no significant group differences in psychiatric diagnoses (**Table 4**) or reversal learning performance changes (**Table 5**).

### Relapse outcome

17 participants relapsed (days to relapse since post-intervention MRI session: M=15.36, SD=10.49) and 43 participants remained abstinent by 1-month. By the 4-month follow-up timepoint 4 participants were not reachable anymore. 25 participants relapsed and 31 remained abstinent by the 4-month timepoint (days to relapse M=44.2, SD=42.42).

### Intervention increased causal connectivity from LDLPFC to specific addiction networks

There was a significant Group (Active tDCS vs. Sham) x Time interaction in the average strength of connectivity from LDLPFC to the IS (LDLPFC-IS; F(1,57)=5.802, p=0.019, **Figure 1a**) and NE (LDLPFC-NE; F(1,57)=7.059, p=0.010) networks (**Figure 1b**). To further examine intervention effects on connectivity change, an analysis of variance (ANOVA) with change in average strength of connectivity as a dependent variable and Group as the independent variable correcting for baseline was conducted. There was a significant Group effect with an increase in LDLPFC-IS (**Figure 2a**) and LDLPFC-NE (**Figure 2b**) causal connectivity in the active tDCS group and a decrease in the sham group. LDLPFC-EF/Go and LDLPFC-EF/Stop causal connectivity did not show a Group effect.

**Figure 1.**
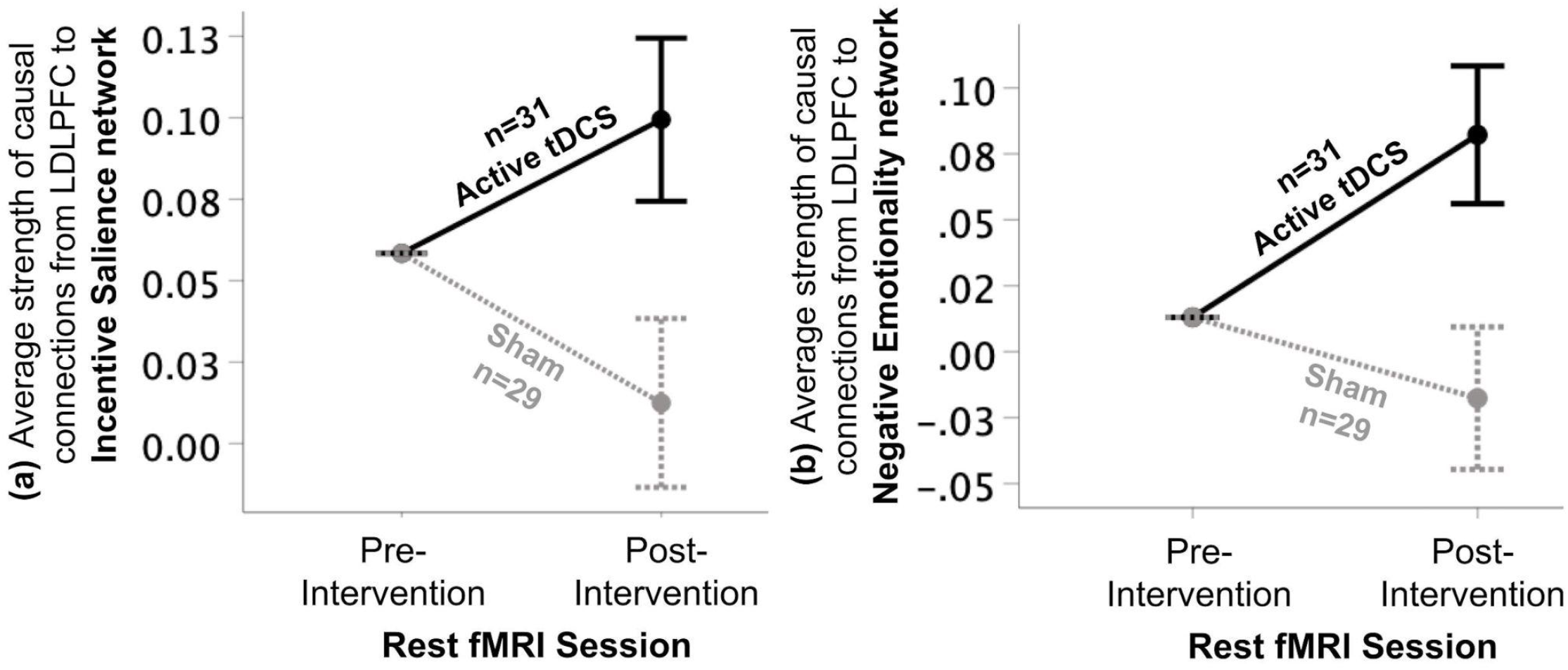
Intervention effects on causal connectivity strength. Repeated measures general linear model analysis correcting for baseline showed a group (active tDCS vs. Sham) x time (pre-vs. post-intervention) interaction effect of average causal connectivity strength from LDLPFC (active stimulation site) to the: **(a)** incentive salience (p=0.019, partial eta2=0.092) and **(b)** negative emotionality (p=0.010, partial eta2=0.110) addiction networks. Error bars: ±1 standard error. tDCS, transcranial direct current stimulation; LDLPFC, left dorsolateral prefrontal cortex; fMRI, functional magnetic resonance imaging.

**Figure 2.**
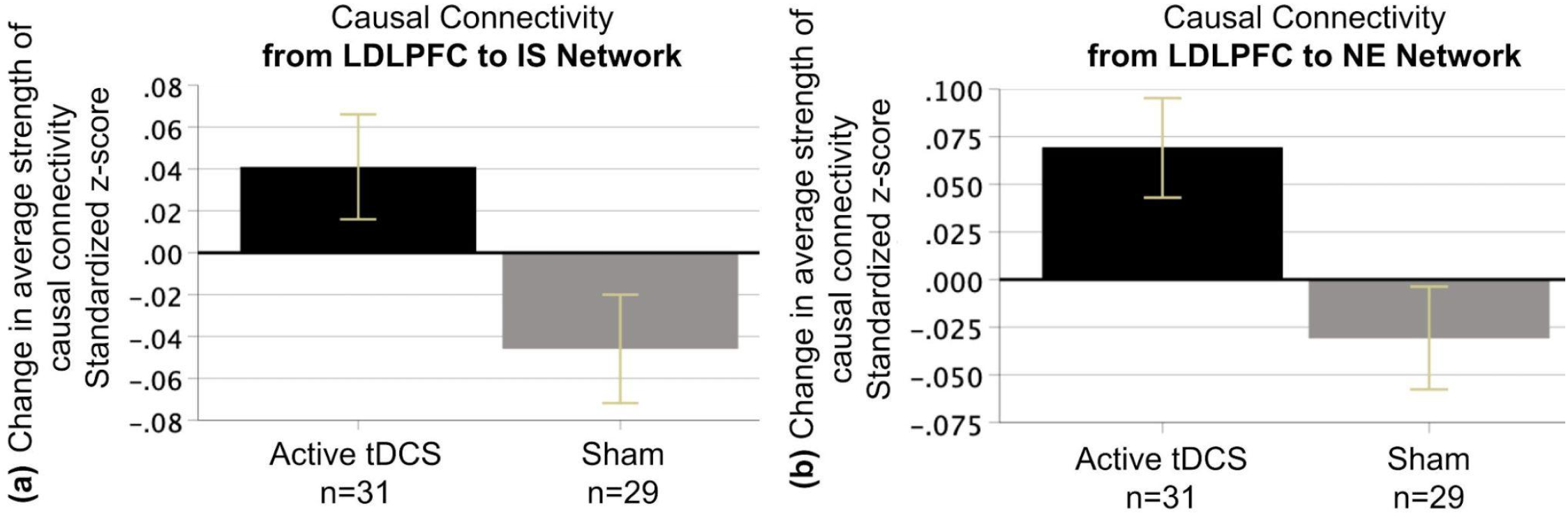
Intervention effects on change in causal connectivity strength. Significant effect of group (Active tDCS vs. Sham) in which those assigned to the active tDCS group showed an increase in causal connectivity strength from LDLPFC to **(a)** incentive salience (F(1,57)=5.802, p=0.019) and **(b)** negative emotionality (F(1,57)=7.059, p=0.010) addiction networks. Those randomly assigned to the sham group showed a decrease in causal connectivity. Error bars: ±1 standard error. tDCS, transcranial direct current stimulation; LDLPFC, left dorsolateral prefrontal cortex; IS, incentive salience; NE, negative emotionality.

### Causal connectivity change predicted 1-month relapse status

Logistic regression with relapse outcome as the dependent variable showed that LDLPFC-IS causal connectivity change was associated with a statistically significant increase in the odds (Odds Ratio=4.15; p=0.015) of remaining abstinent during the 1-month follow-up period (*χ*^2^=7.152, p=0.007). A general linear model analysis correcting for baseline showed a Group (Abstainers vs. Relapsers) x Time interaction effect when examining LDLPFC-IS (F(1,53)=4.230, p=0.045, **Figure 3a**) and LDLPFC-NE (F(1,53)=4.978, p=0.030, **Figure 3b**) causal connectivity strength. An ANOVA with Group (Abstainers vs. Relapsers at the 1-month follow-up period) as the independent variable and causal connectivity change as a dependent variable -correcting for baseline-confirmed a significant group difference in the LDLPFC-IS (**Figure 4a**) and LDLPFC-NE (**Figure 4b**) networks. The effect was characterized by increased connectivity in the active tDCS group and a decrease in the sham group. There were no significant findings with relapse status defined at the 4-month timepoint, or in the EF/Go and EF/Stop networks.

**Figure 3.**
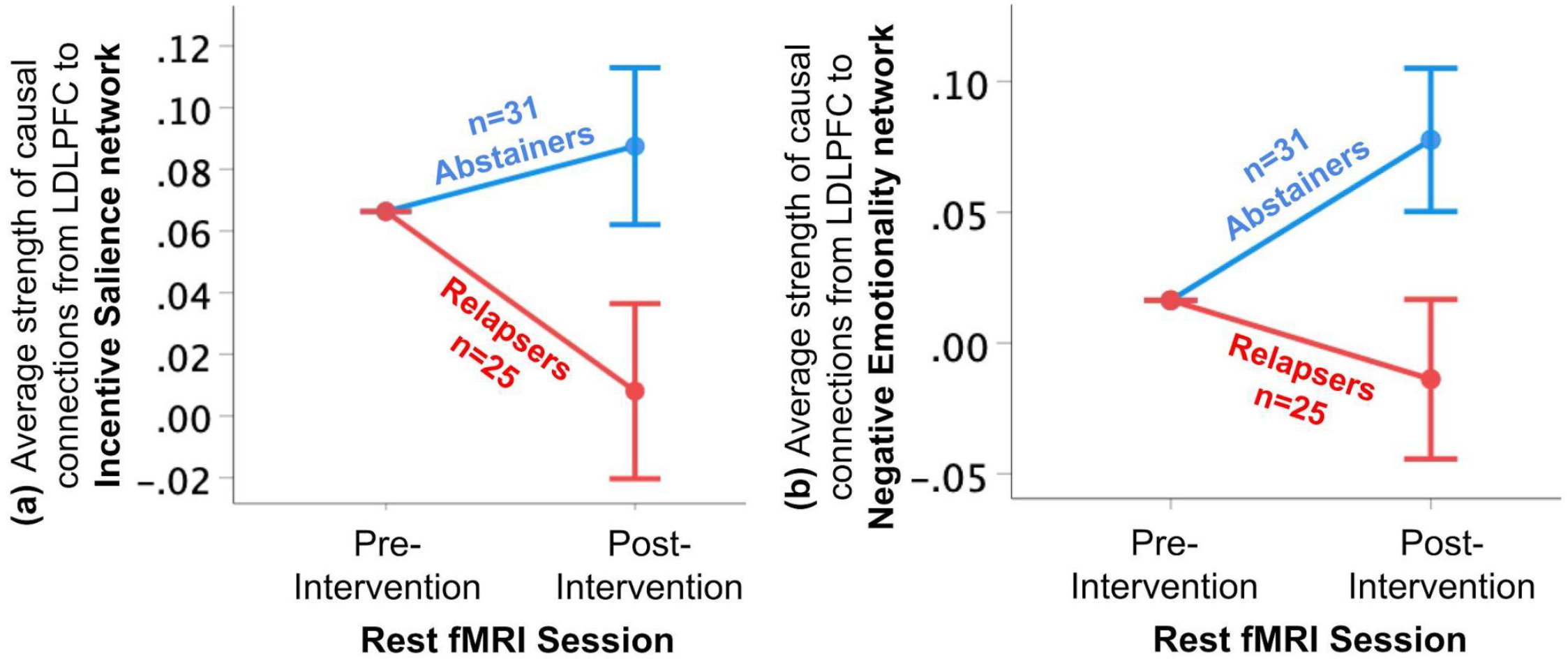
Causal connectivity changes by subsequent relapse status at the 4-month follow-up timepoint. Repeated measures general linear model analysis correcting for baseline showed a group (Abstainers in blue vs. Relapsers in red) x time (pre-vs. post-intervention) interaction effect of average strength of connections from LDLPFC to the **(a)** incentive salience (p=0.045, partial eta2=0.074) and **(b)** negative emotionality (p=0.030, partial eta2=0.086) addiction networks. Error bars: ±1 SE. tDCS, transcranial direct current stimulation; LDLPFC, left dorsolateral prefrontal cortex; fMRI, functional magnetic resonance imaging.

**Figure 4.**
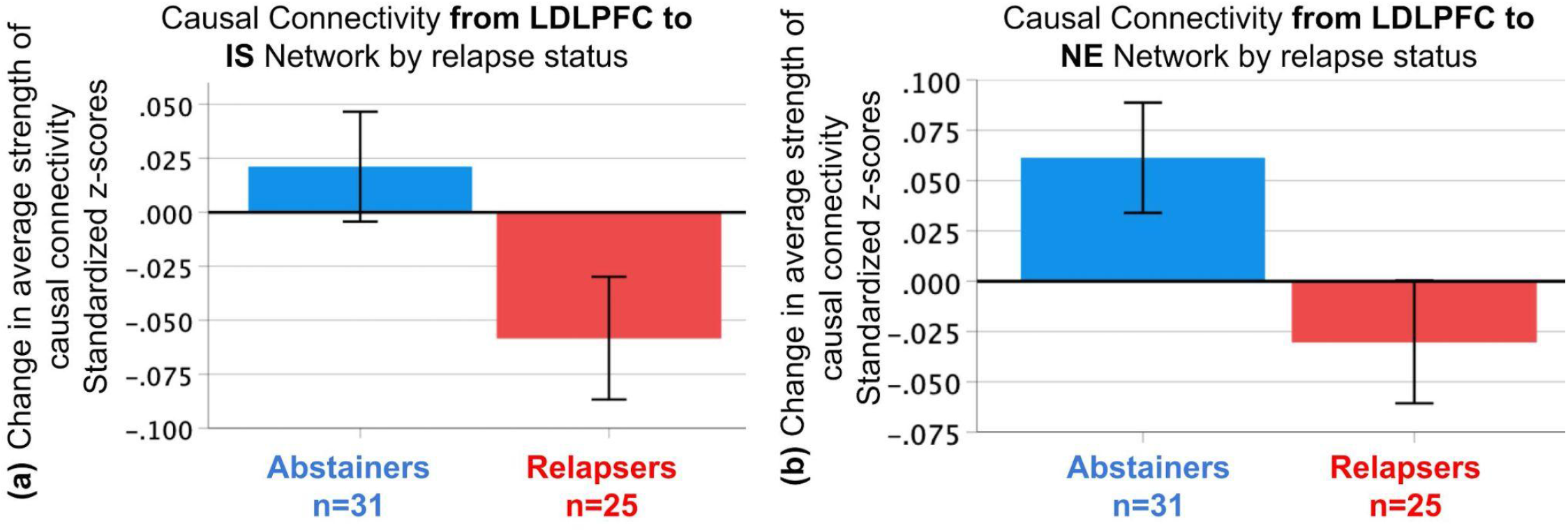
Change in causal connectivity strength by subsequent relapse outcome. Significant effect of group (subsequent Abstainers in blue vs. Relapsers in red) in which those who maintained abstinence over the 4-month follow-up period (Abstainers - blue bar) showed an increase in average causal connectivity strength from LDLPFC to the **(a)** incentive salience (F(1,57)=4.230, p=0.045) and **(b)** negative emotionality (F(1,57)=4.978, p=0.030) addiction networks. Those who relapsed over the 4-month follow-up period (Relapsers - red bar) showed a decrease in causal connectivity. Error bars: ±1 standard error. LDLPFC, left dorsolateral prefrontal cortex; IS, incentive salience; NE, negative emotionality.

### Intervention effects on relapse outcomes

Pearson Chi square revealed that the probability of relapsing in the active tDCS group (n=31) was cut by half when compared to the sham group (n=29) (**Figure 5**).

**Figure 5.**
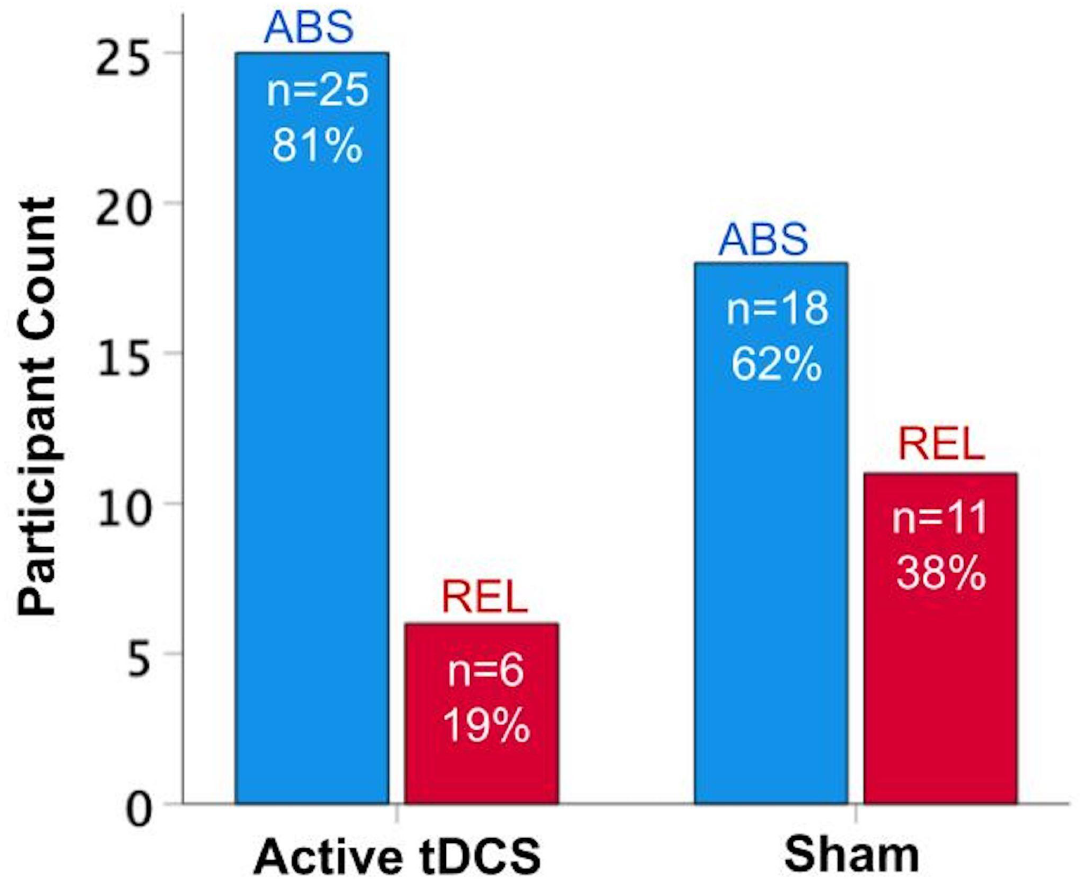
Intervention effects on relapse rates. The probability of relapse in those randomly assigned to receive active tDCS (n=31) was cut by half (19% probability) when compared to those randomly assigned to receive sham (n=29; 38% probability) (Pearson Chi Square=2.55; p=0.11; Phi effect size=0.21).

Because relapse rates were different between men and women (**Supplementary Material F**), we examined whether there were sex dependent intervention effects on relapse outcomes by splitting the sample by sex. There was a significant intervention effect on relapse rates in women (**Figure 6a**), so that the probability of relapsing in women who received active tDCS (n=11) was cut by a factor of 5 when compared to women who received sham (n=10). Men (n=39) did not show a significant effect (**Figure 6b**).

**Figure 6.**
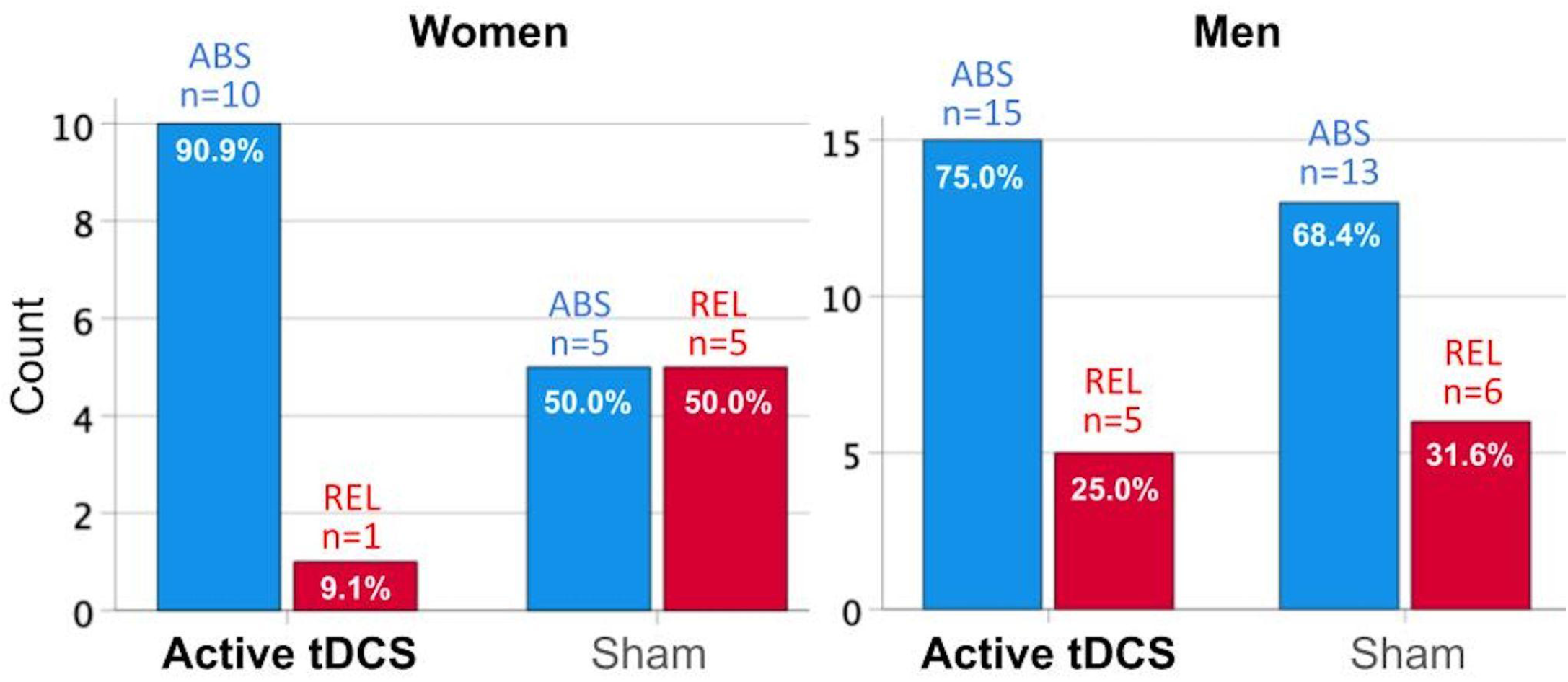
Intervention Effects on Relapse Outcomes by Sex as a Biological Variable. Pearson Chi square analysis with the sample split by sex as a biological variable. **(a)** Women showed significant intervention effects on relapse rates (Pearson Chi Square=4.30; p=0.038; Phi effect size=0.452), while **(b)** men did not (Pearson Chi Square=0.21; p=0.648; Phi effect size=0.073).

### Regression model integrating intervention, LDLPFC-IS causal connectivity change and relapse outcome

A regression model including the main effect of the intervention (Active tDCS vs. Sham), main effect of LDLPFC-IS causal connectivity change, and an interaction between these main effects, showed the following results. First, the main effect of the intervention was associated with the 1-month relapse outcome, so that active tDCS was associated with decreased odds for relapse after adjusting for LDLPFC-IS causal connectivity change and Sex (p=0.012), while sham was not (p=0.227). Second, there was an interaction between connectivity change and intervention group on relapse status during the 1-month follow-up period (interaction coefficient=1.37, p=0.044). Specifically, in the active tDCS group an increase in connectivity change was associated with an increase in the odds of 1-month abstinence, whereas in the sham group a decrease in connectivity change was associated with an increase in the odds of 1-month relapse. The same analysis was not significant for the NE, EF/Go or EF/Stop addiction networks.

### tDCS Side Effects Questionnaire

Average ratings on the questionnaire were <1 (mild) for all symptoms at each timepoint. There were no significant changes from pre-to post-tDCS session, and there were no differences between active tDCS and sham groups for any symptom.

## DISCUSSION

The present study was designed to address the critical need for brain-based interventions that reduce relapse in alcohol use disorder (AUD). We investigated whether a non-invasive neuromodulation intervention combining transcranial direct current stimulation (tDCS) and cognitive training had an effect both on brain networks known to support abstinence and on subsequent relapse rates. While previous studies have reported tDCS effects on brain functional connectivity or clinical outcomes in AUD separately, the present study adds to the literature by providing important converging evidence under one clinical trial. Our results showed that the change in average strength of causal connectivity (from LDLPFC to IS) as a result of active tDCS explains subsequent relapse outcome. The present study further adds to the literature (i) by using recently developed analysis methodologies -causal connectivity analysis-to determine the directional influence of identified connections and (ii) by providing evidence of potential sex-dependent intervention effects on relapse rates.

### tDCS induced causal connectivity changes

Current findings provide two important pieces of evidence regarding tDCS effects on resting state functional connectivity (RSFC). First, our results show that the tDCS intervention modulated RSFC of addiction networks that we previously found to support abstinence [16]. Neuroimaging data collected before and after the intervention allowed us to identify connections being modulated with tDCS. Present findings extend a previous report of tDCS’s effects on global efficiency in AUD [2] by focusing the connectivity analysis from the LDLPFC to specific addiction networks known to support abstinence [16]. We found that active tDCS enhanced the average strength of connections originating from the stimulation area (LDLPFC) to two specific addiction networks: the incentive salience (IS) and the negative emotionality (NE) networks (**Figure 1**). Given that both the IS and NE addiction networks include subcortical/limbic regions (**Table 3**), present findings suggest that LDLPFC stimulation impacts the RSFC of downstream addiction networks previously found to support abstinence [16].

The biochemical mechanisms underlying DLPFC stimulation using tDCS have been previously demonstrated using positron emission tomography (PET) imaging, with important evidence that tDCS on DLPFC induces neurotransmitter release in subcortical areas, that is, increased extracellular dopamine of the striatum [49]. Second, causal connectivity analysis allowed us to determine the direction of identified associations. Causal models showed that LDLPFC stimulation had direct influence on within-network RSFC in the IS and NE addiction networks. These results are promising evidence of the modulatory effects of non-invasive neuromodulation on top-down RSFC, specifically from a key executive control cortical region (DLPFC) to key addiction networks involved in reward processing and emotion regulation [50–52].

### tDCS effects on clinical measures in addiction

The present study extends our most recent cross-sectional neuroimaging findings [16] which reported that higher resting state functional connectivity (RSFC) within the incentive salience (IS), negative emotionality (NE), and executive functioning addiction networks measured during early abstinence was associated with reduced relapse rates and longer periods of abstinence. Our regression model including intervention, connectivity change, and relapse outcome showed that the specific increase in LDLPFC-IS causal connectivity after 5 days of active tDCS was associated with increased odds of maintaining abstinence in the subsequent follow-up. The specificity of the IS addiction network is consistent with our previous report [16] in which within-network IS RSFC had the largest predictive power of subsequent relapse and time to relapse in AUD. Since the IS network consists mainly of striatal regions (**Table 3**) involved in reward processes (e.g. nucleus accumbens, caudate, putamen), current findings are consistent with previous reports pointing to the importance of enhanced engagement of executive control and reward processing networks (i.e. frontal-striatal RSFC) needed for successful recovery [5,6,53].

Our findings also extend existing converging reports of promising effects of frontal (i.e. DLPFC) stimulation in reducing relapse [1,3,17,19]. Previous studies have reported tDCS effects on relapse outcomes over various lengths of follow-up periods, such as over a 2-week [3] or 5-week follow-up period [19]. We are the first tDCS study that completed both a 1-month and 4-month follow-up timepoints. We found that the effect of our intervention on relapse rates was only evident when examining the 1-month follow-up timepoint. This may point to the potential decay of the effect of the intervention beyond 1 month, suggesting the potential need of either increasing stimulation dose (e.g. more intervention days) or the addition of booster sessions. New clinical trials designed to examine the durability of tDCS intervention in AUD need to be conducted.

### Considerations

Future trials designed to specifically address the following issues need to be conducted. First, while the statistical model comparing relapse rates between groups (active-tDCS vs. sham) did show a moderate effect in the whole sample (**Figure 5**), this effect seemed to be driven by women (**Figure 6**). Existing literature suggests that sex-dependent differences may be due to inherent anatomical differences mediating tDCS-induced neuroplasticity. A computational study specifically addressing sex-dependent tDCS effects found 10-11% higher average delivered electric field in women than men when factoring in white/gray matter and cerebral spinal fluid [54]. A tDCS study investigating intervention effects on a task requiring inhibitory control (i.e. Stop-signal) suggested their sex-dependent findings were specific to neural substrates underlying response execution with better intervention-related performance in women than men [55]. Because neuroanatomical and physiological sex differences may underlie sex-dependent tDCS effects, sufficiently powered studies designed to examine the delivered electric field induced by tDCS associated with differences in gray and white matter distribution across sexes need to be conducted. Second, our results suggest that cognitive training with the reversal task alone (sham condition) was not sufficient to induce effects on connectivity change or treatment outcome. A recent study found the highest intervention effect on relapse rates when combining active tDCS with a task requiring inhibitory control (i.e. Go/No-Go task) [3]. Growing evidence suggests that tDCS effects are maximized if delivered concurrently with (i) a variety of cognitive training tasks demanding engagement of different executive function domains (e.g. cognitive flexibility, inhibition, working memory, decision making) and (ii) tasks that continually challenge the participant’s individual ability.

### Conclusion

Results from our longitudinal double-blind randomized clinical trial suggest that 5 days of LDLPFC stimulation delivered during early abstinence (i) increased resting state connectivity of addiction networks supporting abstinence -incentive salience and negative emotionality- and (ii) increases the odds of maintaining abstinence in AUD. More specifically, an increase in LDLPFC-IS connectivity after active stimulation was associated with increased odds of abstinence maintenance in the subsequent month. The unexpected sex-dependent neuromodulation effects need to be further examined in larger clinical trials.

## Data Availability

All data produced in the present study are available upon reasonable request to the authors

## CRediT author statement

**Jazmin Camchong:** Conceptualization, Methodology, Validation, Formal Analysis, Investigation, Writing - Original Draft, Reviewing and Editing, Visualization, Project Administration, Funding Acquisition. **Mark Fiecas**: Methodology, Validation, Formal Analysis, Writing - Original Draft, Reviewing and Editing. **Casey S. Gilmore:** Writing - Original Draft, Reviewing and Editing. **Matt Kushner:** Writing - Reviewing and Editing. **Erich Kummerfeld**: Methodology, Resources (Analysis tools), Writing - Reviewing and Editing. **Bryon A. Mueller:** Data Curation, Writing - Reviewing and Editing. **Donovan Roediger:** Methodology, Formal Analysis, Data Curation. **Kelvin O. Lim:** Conceptualization, Software, Resources (Analysis tools), Writing - Reviewing and Editing.

## ACKNOWLEDGEMENTS

This work was supported by the National Institute of Health (K01AA026349 to M.F and J.C.; UL1TR002494 to J.C.; MH116987 to M.F. and K.O.L.; UG3DA048508 and R01DA038984 to K.O.L.; UL1TR000114 to EK; P41EB027061, P30NS076408, S10OD017974-01 to CMRR) and the Westlake Wells Foundation to J.C.. The opinions and assertions expressed herein are those of the authors and do not necessarily reflect the official policy or position of the National Institute of Health or the Westlake Wells Foundation. All authors have declared that there are no competing or potential conflicts of interest.

## SUPPLEMENTARY MATERIAL

### A. Inclusion and exclusion criteria for all participants

Inclusion criteria were: (i) being enrolled in the inpatient addiction treatment program, (ii) being able to provide written consent and comply with protocol procedures; (iii) meeting DSM-IV lifetime criteria for alcohol dependence [1]. Exclusion criteria were: (i) Any medical condition or treatment with neurological sequelae (i.e. stroke, tumor, loss of consciousness>30 min, HIV); (ii) a head injury resulting in a skull fracture or a loss of consciousness exceeding 30 minutes; (iii) any contraindications for MRI scanning (metal implants, pacemakers or any other implanted electrical device, injury with metal, braces, dental implants, non-removable body piercings, pregnancy, breathing or moving disorder); (iv) any primary psychotic disorder (e.g. schizophrenia, schizoaffective disorder); (v) presence of a condition that would render protocol measures difficult or impossible to administer or interpret; (vi) age outside the range of 18 to 65; (vii) *primary* current substance use disorder diagnosis of a substance other than alcohol except for caffeine or nicotine; (viii) clinical evidence for Wernicke-Korsakoff syndrome; (ix) left handedness, (x) entrance to the treatment program under a court mandate. Nicotine use was not an exclusion criterion but was recorded (**Table 2**). Subjects were permitted to have current comorbid drug use (**Table 2**), but their primary substance use disorder diagnosis needed to be based on alcohol use.

### B. Four-choice reversal learning set-shifting task

In the 4-choice task, participants were presented with four identical rectangles placed along a horizontal axis on the center of the screen. Participants were instructed to select the stimulus that was in the correct location by pressing a button corresponding to its location on the screen. Each of the four rectangle locations had an equal probability of being the correct stimulus choice. Immediate visual feedback for a correct (a check mark) or incorrect (an X) choice was provided. Requirements to change the response set were manipulated by changing the correct location after a variable number of consecutive correct responses (between four and six). This variable number of consecutive correct choices served to reduce the predictability of a reversal and the predictability of receiving negative feedback on a given trial. Each trial (including presentation of stimulus, participant response, and feedback presentation) lasted for 2.5 s, followed by a 500 ms inter-trial interval during which a blank screen was presented. 180 trials were presented over a fixed task duration of 9 minutes.

### C. Brain imaging acquisition and preprocessing

#### Imaging Data Acquisition

Participants underwent two identical MRI sessions - one at pre-intervention and one at post-intervention. All MRI data were acquired from a 3T Siemens Prisma scanner at the University of Minnesota’s Center for Magnetic Resonance Research (CMRR). Acquisition parameters closely matched those created by the Human Connectome Project (HCP)[48–50]. Images collected: a T1-weighted MPRAGE image [TR=2400ms, TE=2.24ms, slices=208, voxel size=0.8mm3], a T2-weighted SPACE image [TR=3200ms, TE=564ms, slices=208, voxel size=0.8mm3], a pair of opposite phase encoded spin echo EPI scans matched to the resting state fMRI, and a resting state fMRI scan [TR=800ms, TE=37ms, slices=72, volumes=520, voxel size=2.0mm3, multiband factor=8, 7 minutes duration]. During the resting state fMRI scan the participant was asked to keep their eyes open, look directly at a fixation cross, to not think of anything in particular, and remain awake (confirmed by participant self-report at the end of each MRI session).

#### Individual anatomical MRI data processing

T1-weighted and T2-weighted images were processed using the HCP minimal preprocessing pipeline [48] completing the following steps: Aligned the T1-weighted and T2-weighted to each other; Registered T1 and T2 images to standard MNI space; Corrected gradient inhomogeneities with gradient distortion correction; Projected images to surface space with FreeSurfer; Motion correction; Removed non-brain tissue; Segmented gray and white matter; Intensity normalization; Extracted and inflated the cortical surface; Output to CIFTI space by extracting NIFTI volumes containing subcortical structural and GIFTI surface files corresponding to the left and right cortical hemispheres, respectively [48].

#### Individual resting state data quality assessment

Resting state fMRI data quality was assessed using methods similar to those outlined in Power et al [51]. Framewise Displacement (FD) and DVARS (“D”: temporal derivative of time courses, “VARS”: root means square -RMS-variance over voxels) were calculated on the resting state fMRI scans. FD is a measure of the head position change relative to the previous time point. DVARS is a measure of the RMS signal change from the previous time point, calculated over the brain mask [51]. Volumes with FD>0.5mm and/or DVARS>0.008 were flagged as bad along with the previous volume and next 2 volumes. Resting scans with more than 30% flagged volumes were rejected from the study. Three participants were excluded from analyses because they failed this quality criteria (see Participants section).

#### Individual fMRI data preprocessing

After individual anatomical MRI data processing, the following preprocessing steps (similar to [15]) were completed in chronological order on resting state fMRI data. First, nonlinear distortions produced by the gradient were corrected using the HCP version of the gradunwarp package 1. Second, EPI (echo-planar imaging) realignment to correct for motion by registering each volume to the single band reference image via FLIRT with 6 degrees of freedom. To correct for distortion in the phase encoding direction, a pair of opposing phase encoding spin echo EPI field maps were used to estimate a distortion field with the FSL tool “topup” and applied to the EPI fMRI images with FLIRT. Following distortion correction, EPI to T1-weighted surface registration and native to MNI nonlinear registration were performed. Finally, the voxels of the MNI registered volumes were resampled onto cortical surfaces and extracted to CIFTI space.

#### Individual functional processing

Because the rest fMRI analysis relied on temporal correlations, the primary focus of the functional pipeline was to correct temporal artifacts. First, to linearly detrend the data, a high band-pass filter cutoff of 2000 secs with a slow roll off [49] was applied. Second, the data was denoised using ICAFIX [52,53] with a customized FIX classifier using 15 randomly selected scanning sessions from the current AUD data. This customized FIX classifier was trained by J. Camchong -ICAFIX Pipeline Documentation-to apply a classifier with “noise” components characteristic to an AUD sample (e.g. larger ventricles). “FIX” classified the spatial components as “signal” or “noise”. Components classified as noise were regressed out from the data (multiplied by the associated time series and subtracted from the original dataset) [49]. Following the ICAFIX process, surface-based functional alignment was run on the ICAFIX denoised data with the tool MSMAll [54]. MSMAll employed myelin maps, resting state fMRI network maps, and resting state fMRI visuotopic maps to align a participant’s cortical data to a group template. Once complete, the MSMAll and ICAFIX fMRI concatenated data was dissociated resulting in fully preprocessed resting state fMRI data, used for all subsequent analyses.

#### Individual data segmentation

Preprocessed resting state fMRI individual data was parcellated into distinct and non-overlapping regions using the cortical Schaefer 400 (i.e. 400 regions across the cortex)[55] and the subcortical Harvard-Oxford atlases [56]. Within each region, the individual grayordinates [57] were averaged together per time point to generate a parcellated time series of the resting state fMRI data.

#### Individual causal connectivity analyses

We implemented and conducted computations on the parcellated time series. We used causal discovery analysis (CDA) to generate individual causal connectomes of left dorsolateral prefrontal cortex (LDLPFC, tDCS stimulation site) and theoretically defined addiction networks ([2,3], **Table 3**). The analysis workflow proceeded as follows. First, for each participant, we selected the Schaefer parcellations [4] to define ROIs that corresponded to the LDLPFC and the theoretically defined addiction networks: the incentive salience (IS), executive functioning-Go (EFG), executive functioning-Stop (EFS) and negative emotionality (NE) networks (As in [3]). Time series data from these selected parcellations (**Table 3**, details in **Supplementary Material D and E**) were extracted. Second, to generate causal models of LDLPFC’s directional influence on addiction networks we used the CDA method Greedy Adjacencies and Non-Gaussian Orientations (GANGO [5]). GANGO allowed us to identify nodal adjacencies [6–8] and edge orientations [9]. This method has been used to generate causal models of AUD [10] and fMRI data [5]. Third, to quantify the strength of each connection we used the GANGO causal models to determine the structure of a structural equation model (SEM), and fit that SEM to the time series data. We then extracted the strength (represented by standardized r score) and sign (positive or negative) of each identified directional edge from the fitted SEM. The strength of the modeled causal relationship from the LDLPFC ROI to each addiction network ROIs was quantified by calculating the average strength of identified directional edges (sum of all edge strengths / number of edges) between the parcels in the pair of ROIs. In the end this produced, for each participant, an averaged standardized correlation value representing the strength of the causal connectivity from LDLPFC to each addiction network.

### D. Schaefer parcels within the left dorsolateral prefrontal cortex (LDLPFC). As in our previous manuscript [3]: Regions within each domain-defined addiction network (Table 3) with corresponding cortical Schaefer [4] or subcortical Harvard-Oxford [56] atlas parcels listed

#### Left Dorsolateral Prefrontal Cortex (LDLPFC)

**Cortical**: ContA_PFCl_1, 2, 3; ContA_PFClv_1, 2; ContB_PFCd_1; ContB_PFClv_1, 2, 3;

DefaultA_PFCd_1, 2, 3; DefaultA_PFCm_1, 3, 4, 5; DefaultB_PFCd_1, 2, 3, 4, 5, 6;

DefaultB_PFCl_1; DefaultB_PFCv_1, 2, 3, 4, 5; LimbicB_OFC_1, 2, 3, 4, 5;

SalVentAttnA_FrOper_2; SalVentAttnB_OFC_1; SalVentAttnB_PFCl_1, 2, 3.

#### Incentive Salience (IS)

**Subcortical:** Caudate, Nucleus Accumbens, Pallidum, Putamen

**Cortical:** Motor cortex corresponding to Schaefer regions: SomMotA_1, 5, 7, 8, 9, 10, 11, 12,

13, 15, 16, 17, 18, 19; SomMotB_Cent_1, 2, 3, 4, 5; DefaultB_PFCl_2; SalVentAttnA_FrMed_2,

3, 4; SalVentAttnA_ParMed_1, SalVentAttnA_PrC_1, ContA_PFCd_1.

#### Negative Emotionality (NE)

**Subcortical:** Amygdala, Caudate, Nucleus Accumbens, Pallidum, Putamen, Ventral Diencephalon.

#### Executive Function GO (EF/Go)

**Subcortical:** Caudate, Nucleus Accumbens, Pallidum, Putamen

**Cortical:** Anterior Cingulate Cortex corresponding to Schaefer regions: SomMotA_6, 13, 14, 18, 20; DorsAttnB_PostC_8, 9; SalVentAttnA_ParMed_1, 2, 3, 4. Inferior Frontal Cortex corresponding to Schaefer regions: SalVentAttnA_FrOper_2; SalVentAttnA_FrOper_3; ContA_PFClv_1; ContA_PFCl_1, 2; DefaultB_PFCv_3, 4, 5; SalVentAttnB_PFClv_1. Insula corresponding to Schaefer regions: SalVentAttnA_Ins_1, 2, 3, 4; SalVentAttnB_Ins_1, 2, 3; SalVentAttnA_FrOper_1; SomMotB_Ins_1; SomMotB_S2_1, 2, 4, 6, 7. Medial Prefrontal Cortex corresponding to Schaefer regions: SalVentAttnA_FrMed_1; ContA_Cingm_1; ContB_PFCmp_1; DefaultA_PFCm_1, 2, 3, 4, 6; DefaultB_PFCd_1, 3; SalVentAttnB_PFCmp_1, 2; LimbicB_OFC_3.

#### Executive Function STOP (EF/Stop)

**Subcortical**: Amygdala

**Cortical**: Anterior Cingulate Cortex corresponding to Schaefer regions: ContA_PFCl_3,5; ContA_PFClv_2; ContB_PFCd_1; ContB_PFCld_1,2,3,4; ContB_PFClv_3; DefaultA_PFCd_1,2,3; DefaultA_PFCd_2,3,4,5,6; DefaultA_PFCm_5; DefaultB_PFCl_1; DefaultB_PFCd_1,2,3,4,5,6; SalVentAttnB_PFCl_1,2,3. Dorsolateral Prefrontal Cortex corresponding to Schaefer regions: ContA_PFCl_3,5; ContA_PFClv_2; ContB_PFCd_1; ContB_PFCld_1,2,3,4; ContB_PFClv_3; DefaultA_PFCd_1,2,3; DefaultA_PFCd_2,3,4,5,6; DefaultA_PFCm_5; DefaultB_PFCl_1; DefaultB_PFCd_1,2,3,4,5,6; SalVentAttnB_PFCl_1,2,3. Inferior Frontal Cortex corresponding to Schaefer regions: ContA_PFCl_1,2; ContA_PFClv_1; DefaultB_PFCv_3,4,5; SalVentAttnA_FrOper_2,3; SalVentAttnB_PFClv_1. Medial Prefrontal *Cortex* corresponding to Schaefer regions: ContA_Cingm_1; ContB_PFCmp_1; DefaultA_PFCm_1,2,3,4,6; DefaultB_PFCd_1,3; SalVentAttnA_FrMed_1; SalVentAttnB_PFCmp_1,2. Orbitopolar Frontal Cortex corresponding to Schaefer regions: ContB_PFClv_1, 2, 3, 4; SalVentAttnB_OFC_1; LimbicB_OFC_1, 2, 3, 4, 5, 6; DefaultA_PFCm_3, 5; DefaultB_PFCv_1, 2.

**E**. Figures of theoretically defined addiction networks, same networks as in [3](Table 2 and Supplemental Material D list regions).

**Table 2.** Counts of lifetime and current substance use disorder for all participants with alcohol use disorder.

| **Substance** | **Lifetime Diagnosis Count** |  |  | **Current Diagnoses Count** |  |  |
| --- | --- | --- | --- | --- | --- | --- |
| | Active<br>tDCS<br>(n=31) | Sham<br>(n=29) | $\chi^2$<br>Sig. | Active<br>tDCS<br>(n=31) | Sham<br>(n=29) | $\chi^2$<br>Sig. |
| Marihuana | 12 | 13 | p=0.58 | 7 | 5 | p=0.53 |
| Cocaine | 5 | 4 | p=0.59 | 0 | 0 | -- |
| Methamphetamine | 4 | 4 | p=0.62 | 0 | 0 | -- |
| Opioids | 1 | 2 | p=0.51 | 0 | 0 | -- |
| Hallucinogens | 2 | 4 | p=0.41 | 0 | 0 | -- |
| Nicotine | 17 | 18 | p=0.67 | 17 | 16 | p=0.91 |

tDCS, transcranial direct current stimulation; $\chi^2$ , Chi-Square; p, significance (Sig.) probability value.

**Table 3.**
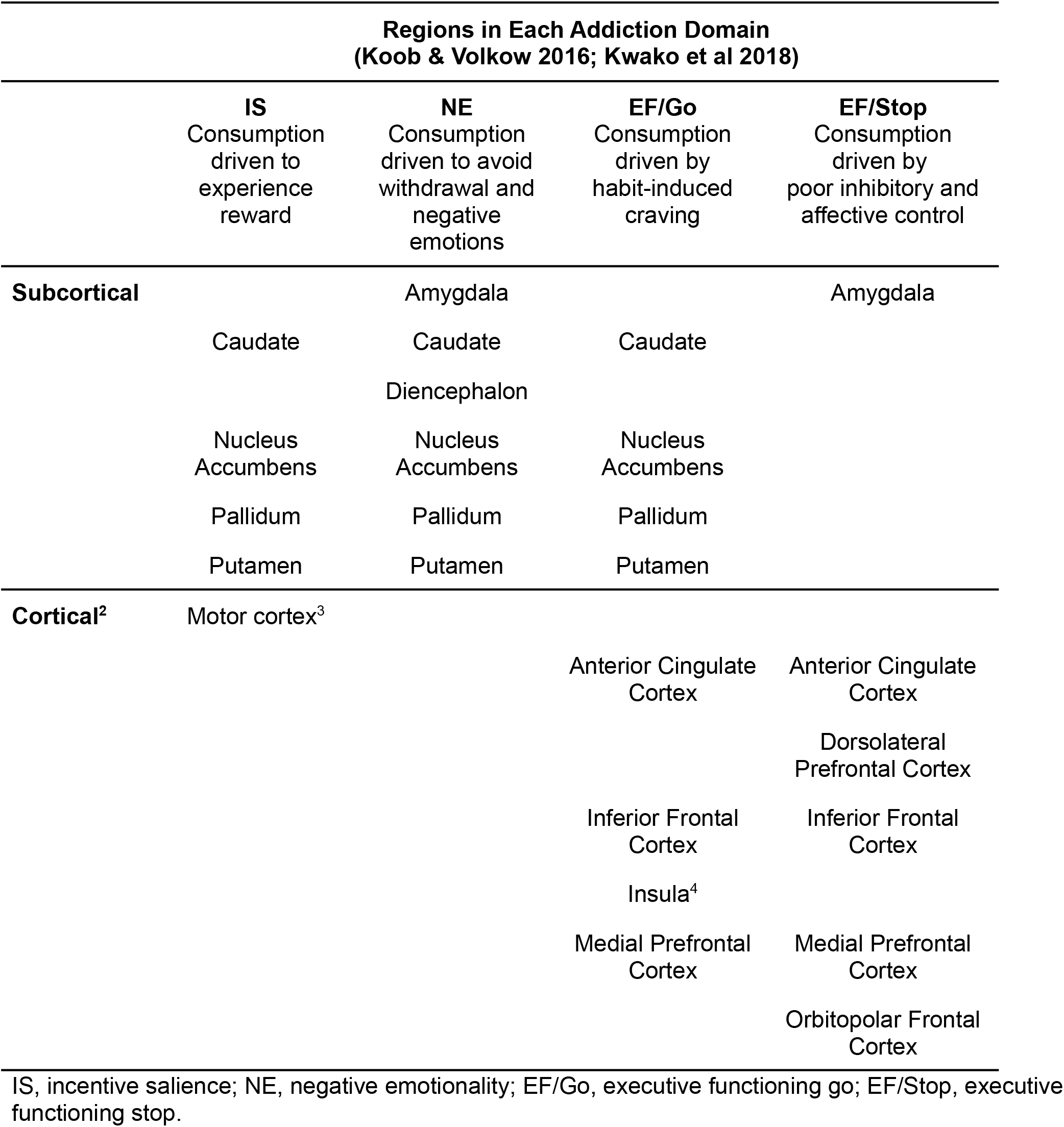
Bilateral brain regions within each Addiction Domain

**Table 4.** Counts of lifetime and current psychiatric diagnoses by intervention group.

| Psychiatric<br>Diagnoses<sup>6</sup> | Lifetime Diagnosis Count (%) |  |  | Current<sup>5</sup> Diagnoses Count (%) |  |  |
| --- | --- | --- | --- | --- | --- | --- |
| | Active | Sham | $\chi^2$ | Active | Sham | $\chi^2$ |
|  | tDCS<br>(n=31) | (n=29) | Sig. | tDCS<br>(n=31) | (n=29) | Sig. |
| MDD | 13 | 14 | p=.62 | 13 | 13 | p=.81 |
| GAD | 14 | 9 | p=.26 | 11 | 9 | p=.72 |
| PTSD | 8 | 11 | p=.31 | 9 | 10 | p=.75 |
| Social phobia | 8 | 7 | p=.89 | 7 | 7 | p=.89 |
| PD | 6 | 7 | p=.65 | 3 | 4 | p=.62 |
| Agoraphobia | 3 | 3 | p=.93 | 2 | 3 | p=.59 |
| ADHD | 3 | 1 | p=.33 | 3 | 1 | p=.33 |

tDCS, transcranial direct current stimulation; $\chi^2$ , Chi-Square; MDD, major depressive disorder; GAD, generalized anxiety disorder; PTSD, post-traumatic stress disorder; PD, panic disorder; ADHD, Attention deficit hyperactivity; p, significance (Sig.) probability value.

**Table 5.** Reversal Learning task performance change (post-intervention minus pre-intervention).

|  | **All AUD** | **Active tDCS** | **Sham** | **Sig.** |
| --- | --- | --- | --- | --- |
| **Performance change** | **(n=60)** | **(n=31)** | **(n=29)** |  |
| Mean reversal trial response<br>time | -0.45 | -0.48 | -0.42 | p=0.79 |
| Number of Reversals | 3.38 | 2.19 | 4.45 | p=0.12 |

AUD, Alcohol Use Disorder; tDCS, transcranial direct current stimulation; p, significance (Sig.) probability value.

### Incentive Salience (IS) addiction network

First two columns are IS regions corresponding to cortical Schaefer 400 atlas parcels on bilateral motor cortex displayed on an MNI surface brain. Last three columns are IS regions corresponding to subcortical Harvard-Oxford atlas parcels on bilateral caudate, nucleus accumbens, pallidum, and putamen on axial slices (z) displayed on an MNI average brain. A, anterior; P, posterior; L, left; R, right.

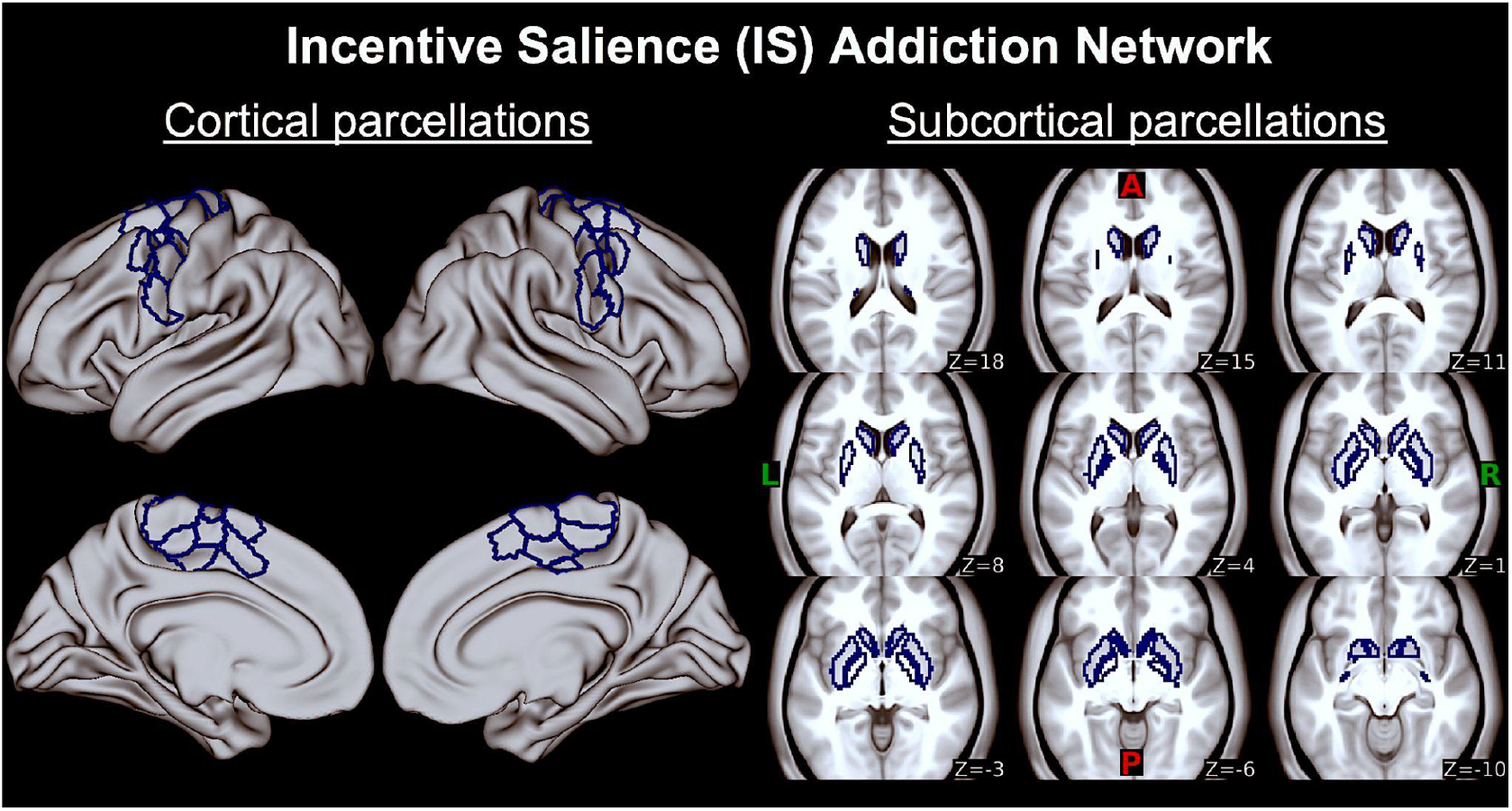

### Negative emotionality (NE) addiction network

Last three columns are NE regions corresponding to subcortical Harvard-Oxford atlas parcels on bilateral amygdala, caudate, habenula, nucleus accumbens, pallidum and putamen on axial slices (z) displayed on an MNI average brain. There are no cortical parcels within this addiction network, so two first columns with the MNI surface brain are empty. A, anterior; P, posterior; L, left; R, right.

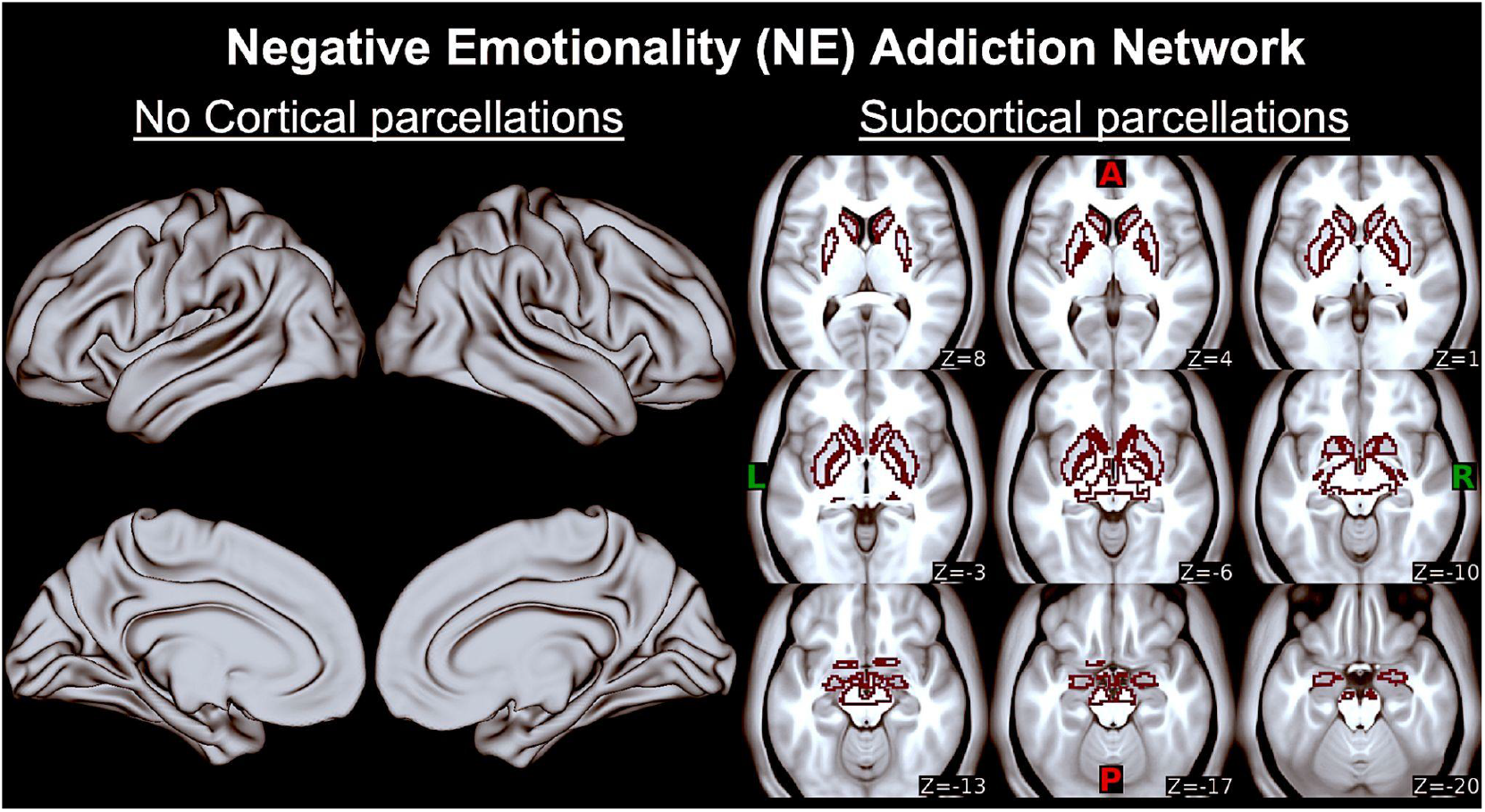

### Executive function Go (EF/Go) addiction network

First two columns are EF/Go regions corresponding to cortical Schaefer 400 atlas parcels on bilateral anterior cingulate cortex, mid cingulate cortex, inferior frontal cortex, insula (anterior agranular insular complex, frontal opercular, insula granular, middle insula, posterior insula, posterior opercular), and medial prefrontal cortex displayed on an MNI surface brain. Last three columns are EF/Go regions corresponding to subcortical Harvard-Oxford atlas parcels on bilateral caudate, nucleus accumbens, pallidum and putamen on axial slices (z) displayed on an MNI average brain. A, anterior; P, posterior; L, left; R, right.

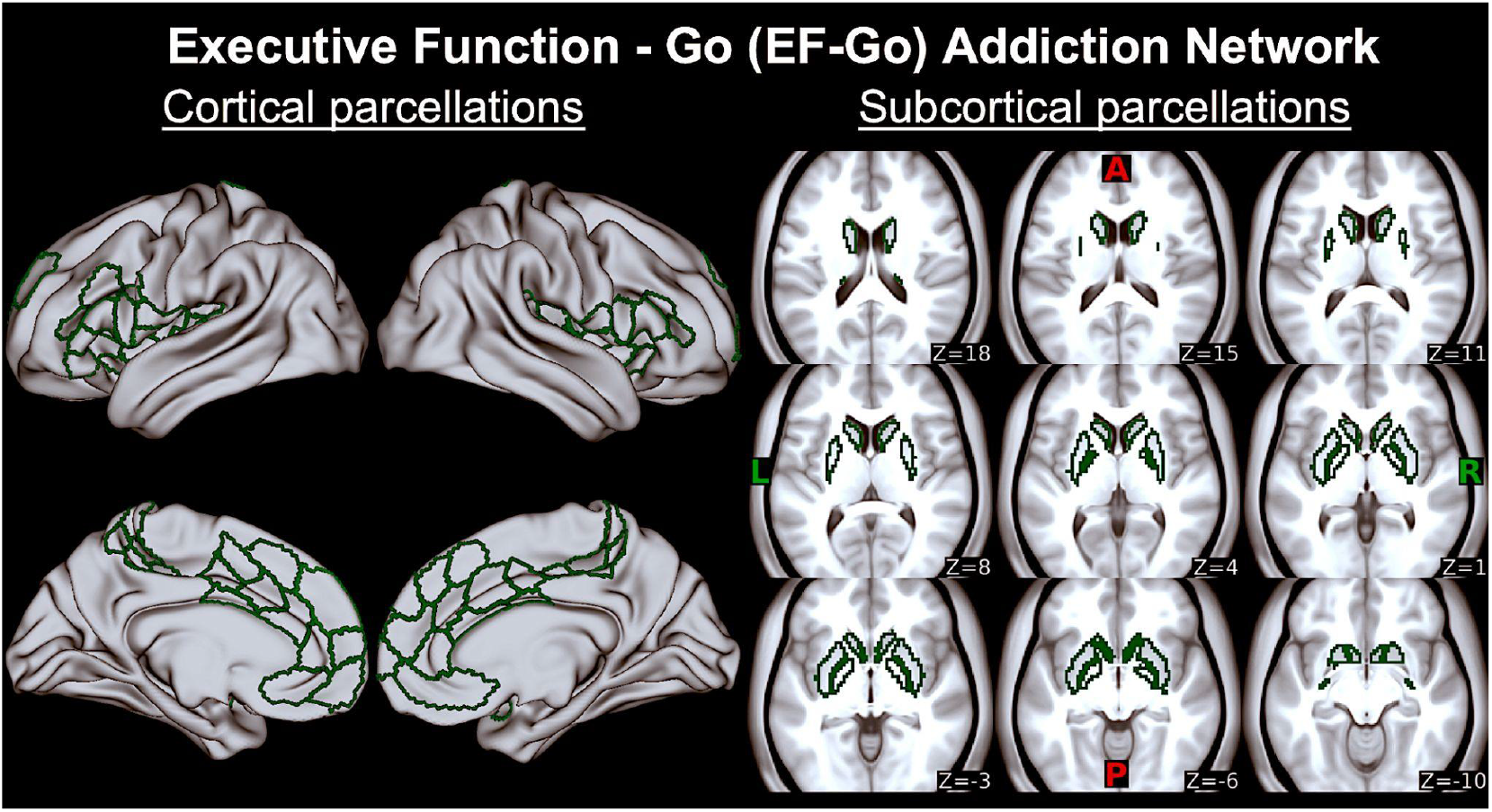

### Executive function Stop (EF/Stop) addiction network

First two columns are EF/Stop regions corresponding to cortical Schaefer 400 atlas parcels on bilateral anterior cingulate cortex, dorsolateral prefrontal cortex (BA 6, 8, 9, 46), inferior frontal cortex, medial prefrontal cortex, orbitopolar frontal cortex (BA 10, 47) displayed on an MNI surface brain. Last three columns are EF/Stop regions corresponding to subcortical Harvard-Oxford atlas parcels on bilateral amygdala on axial slices (z) displayed on an MNI average brain. A, anterior; P, posterior; L, left; R, right.

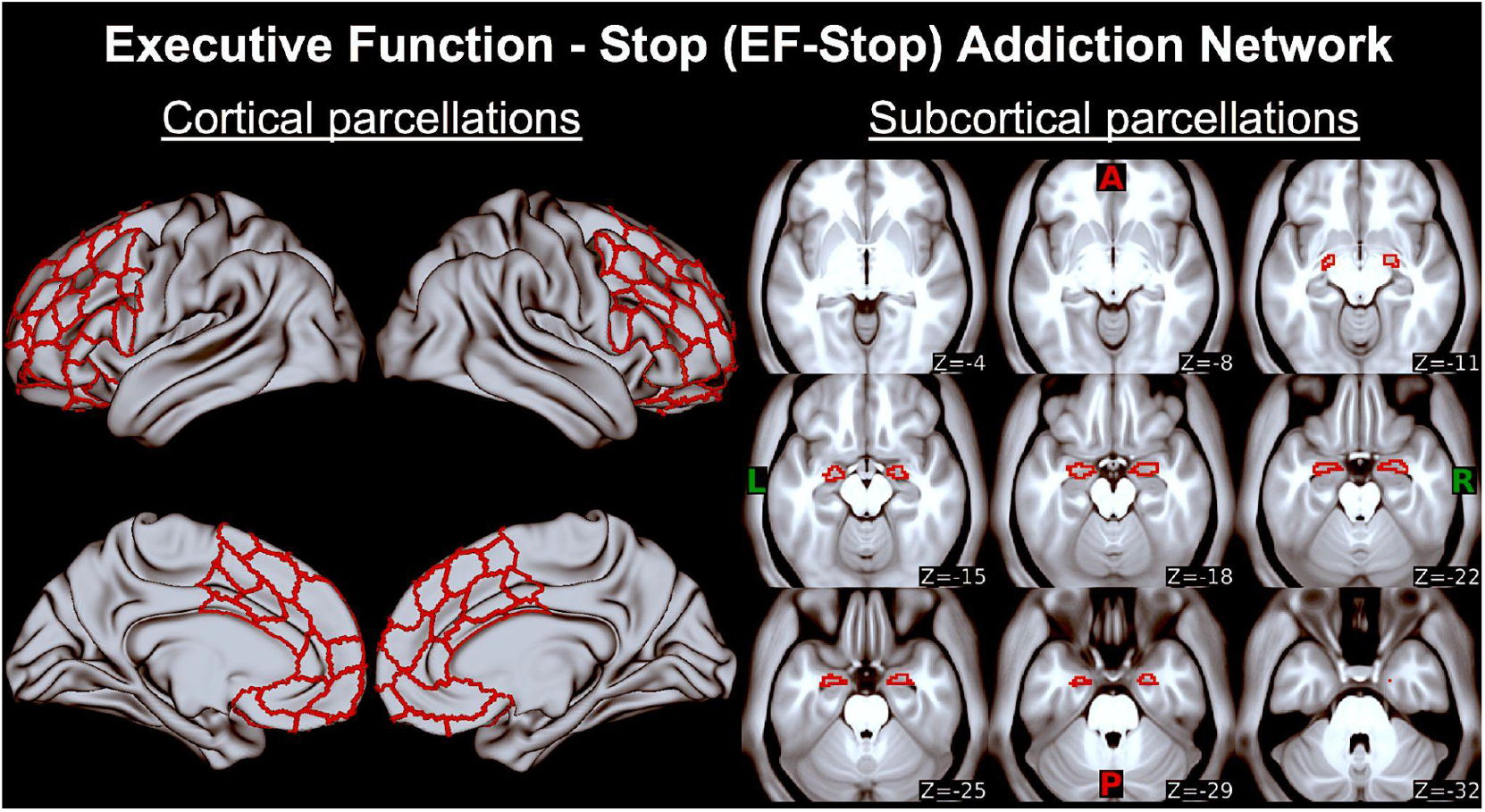

### F. Sex differences in survival rates

Cox’s proportional hazards regression showing cumulative survival curves by sex as a biological variable (collapsed across active tDCS and sham treatment groups) over the 4-month follow-up period. Results suggested that being female increased the odds of relapse (Hazard Ratio=2.427) during the 4-month period [*χ*^2^=4.692, p=0.03].

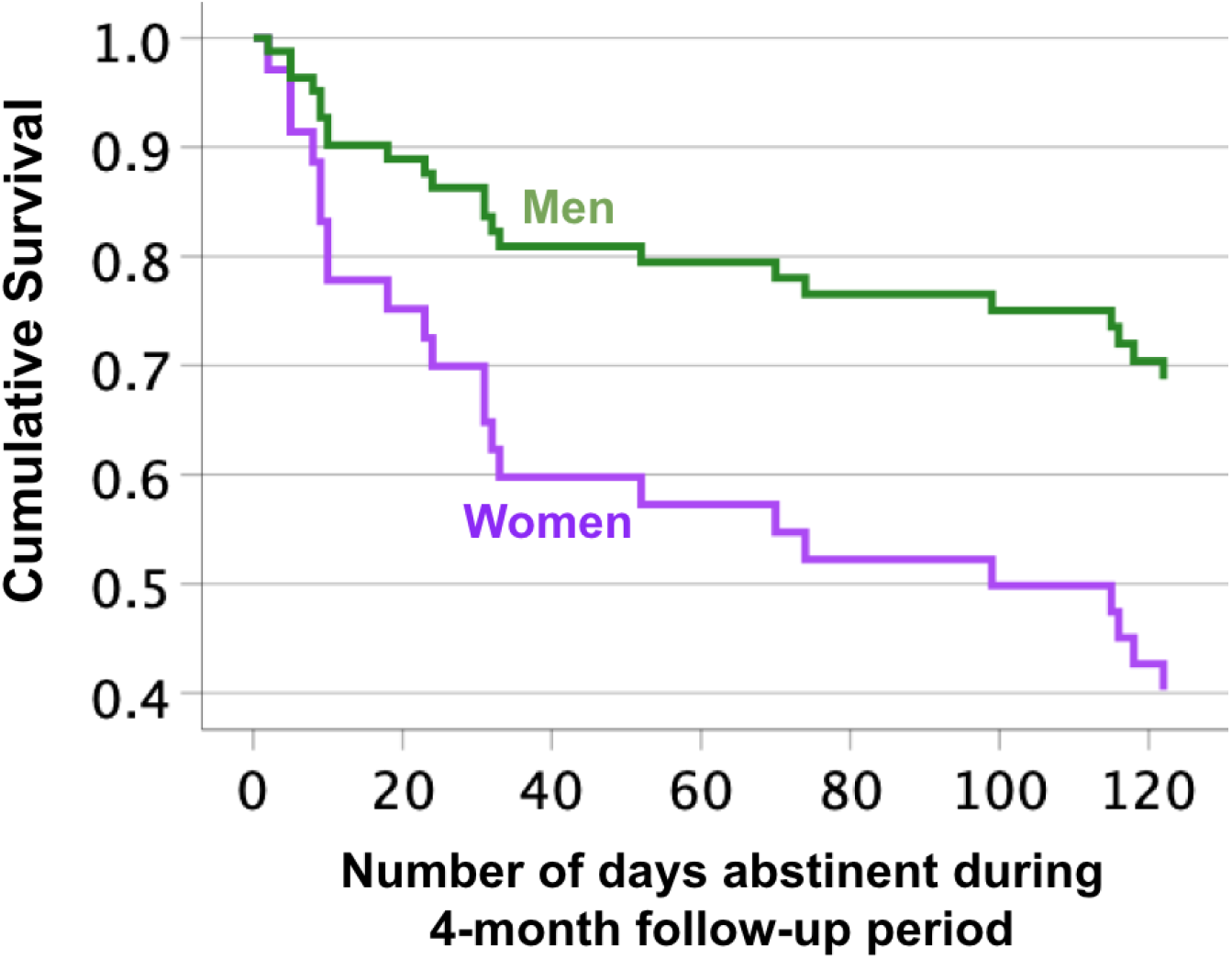

## Footnotes

1 Potential side effects included headache, neck pain, scalp pain, tingling, itching, burning sensation, skin redness, sleepiness/fatigue, poor concentration, acute mood change, and nausea. Severity was rated on a scale: 0 (absent), 1 (mild), 2 (moderate), 3 (severe).

2 Corresponding Schaefer regions in **Supplemental Material D**

3 Including: Inferior and superior premotor, pre-motor, somatosensory, supplementary motor

4 Including: Anterior agranular insular complex, frontal opercular, insula granular, middle insula, posterior insula, posterior opercular

5 Participants with current diagnoses were clinically stable.

6 No lifetime or current diagnoses for the following disorders in the current sample: Dysthimia, Hypomania, Bipolar disorder (without psychosis episodes), Obsessive compulsive disorder, Antisocial personality disorder, Conduct disorder.

